# Selective Outcome Reporting in Cancer Studies: A Scoping Review

**DOI:** 10.1101/2024.07.02.24309826

**Authors:** Jennifer Hinkel, Carl Heneghan, Clare Bankhead

**Affiliations:** University of Oxford

## Abstract

**Background:** Unbiased reporting of clinical study results is essential for evidence-based medicine. However, Selective Outcome Reporting (SOR) leads to Outcome Reporting Bias (ORB) and is prevalent across disease areas, including oncology. This scoping review aims to: (a) describe the current state of research on SOR in cancer studies, (b) assess the prevalence of SOR, (c) understand methods and definitions used in SOR assessment, (d) map available evidence and identify research gaps, and (e) discuss research and policy implications.

**Methods:** A systematic literature search was conducted using keywords related to endpoint discrepancies and oncology. Studies were screened, deduplicated, and evaluated. The JBI Critical Appraisal Checklist for Systematic Reviews and Research Synthesis was used for quality assessment.

**Results:** Six systematic reviews, each including 24 to 217 cancer clinical trials, were analysed. SOR prevalence varied from 4% to 79%, with a median rate of 12%. Definitions of endpoint discrepancies varied, complicating direct comparisons. SOR was identified as over-reporting, under-reporting, or misreporting outcomes.

**Conclusion:** SOR is a significant issue in oncology clinical trials, with implications for evidence synthesis, clinical practice, and policy. The lack of consistent definitions and detailed protocol reporting contributes to the challenge. Enhancing transparency and standardisation in outcome reporting could mitigate ORB and improve the reliability of clinical evidence. Implications: Future research should focus on consistent SOR definitions and improved protocol transparency. Policymakers and regulators should promote standards to reduce SOR and ensure transparent and trustworthy clinical trial outcomes.

## Background

Unbiased reporting of clinical study results is the keystone of evidence-based medicine. Discrepancies in reporting study outcomes exist and have been documented across numerous disease areas and study settings. (Mathieu et al. 2009) Selective Outcome Reporting (SOR) is frequently described as over-reporting, under-reporting, or misreporting and is a source of potential Outcome Reporting Bias (ORB) in clinical study publications. (Kirkham et al. 2018; Thomas and Heneghan 2022)

From an assessment perspective, over-reporting and under-reporting are more easily identified than misreporting. Assessing misreported outcomes requires fully understanding the definitions and measurements proposed in the pre-specified endpoints and the actual measurements used. Both protocols and publications may lack this level of detailed definition and specificity.

Prospective registration of clinical trial protocols in publicly available registries is presently an established standard. Registration is required by medical journals and, in some countries, by statute. In the United States, the US Food and Drug Administration (FDA) Modernization Act established this requirement in 1997; the US National Institutes of Health (NIH) launched clinicaltrials.gov in 2000 as a national public registry of trials. The International Committee of Medical Journal Editors (ICMJE) likewise requires the registration of trials in a public registry. The CONSORT Statement from 1996, states that primary and secondary endpoints should be pre-specified and “completely defined” “including how and when they were assessed.” (Begg et al. 1996) As a result of these mandates and requirements, researchers and practitioners theoretically have access to clinical trial protocols with complete definitions of primary and secondary outcomes. However, protocols may be missing in practice or contain inadequate data to assess all aspects of Selective Outcome Reporting.

While Selective Outcome Reporting presents issues for evidence across disease states and therapeutic areas, some areas present higher stakes. With increasing pressures on healthcare resource utilisation, clinicians seek to apply evidence-based and cost-effective interventions to advance favourable patient outcomes. In recent years, oncology has been in the spotlight, particularly associated with accelerated marketing approvals based on non-traditional trial designs, surrogate endpoints, and Bayesian p-value evaluations. Treatment decisions in oncology represent not only an impact on the patient but also on the health system in terms of resource utilisation and cost. Moreover, within these complex and novel trial designs and statistical methods used to analyse their outputs, specified outcomes are frequently surrogate or proxy endpoints, where measurement and definition may not be as straightforward as final endpoints (for example, Progression Free Survival, PFS, and Overall Response Rate, ORR v. Overall Survival, OS).

To establish a baseline of understanding of the evidence describing Selective Outcome Reporting within the field of oncology, this scoping review analyses published systematic reviews examining endpoint discrepancies in cancer studies. The rationale for positioning this study as a scoping review is that it seeks to identify and characterise what evidence is available and how it is compiled and identify gaps in understanding. These are common purposes of scoping reviews. (Munn et al., 2018).

While framed as a scoping review, this study will also potentially identify and characterise quantitative metrics, including descriptive statistics on the frequency of endpoint discrepancies in cancer studies.

## Methods

The primary objective is to describe the current state of research evidence in outcome reporting and outcome reporting discrepancies/Selective Outcome Reporting (SOR) in cancer.

The secondary objectives are (a) to assess the prevalence of Selective Outcome Reporting in cancer studies according to the existing evidence, (b) to better understand the methods and definitions used to assess SOR in cancer studies, (c) to map the available evidence and identify any potential limitations or gaps in the current research, and (d) to identify and discuss implications for research and policy emerging from these findings.

This review further seeks to explore whether methods to determine Selective Outcome Reporting are defined consistently, whether a common taxonomy exists to describe SOR and related endpoint discrepancies, and whether researchers have found variation in the types of studies with higher or lower rates of Selective Outcome Reporting.

The protocol for this review and subsequent revisions have been published on FigShare. (See: https://figshare.com/account/home#/projects/125815)

### Search strategy and inclusion criteria

The search strategy and inclusion criteria below were defined and published to FigShare. I performed a PubMed search in July of 2022 for papers published between Jan. 1, 2015, and June 30, 2022, with the following keyword phrases and with either “cancer” or “oncology”: “endpoint variability”, “endpoint inconsistency”, “endpoint fidelity,” “endpoint reporting”, “outcome reporting,” “selective reporting of outcomes,” and “reporting discrepancies”. For each keyword phrase searchers, the complete list of resulting citations was downloaded from PubMed and stored in an Excel spreadsheet for review and further screening. Inclusion criteria were for reviews of oncology studies that included quantitative metrics comparing endpoints in a study protocol to endpoints published or reported at the conclusion of a study. An additional search of the same terms as applied to conference papers and abstracts for the major oncology conferences since 2015: ASCO (American Society of Clinical Oncology), AACR (American Association of Cancer Research), and ASH (American Society of Hematology).

Subsequently, each list was reviewed first with a title screen. The included papers were then de-duplicated and reviewed with an abstract screen. Of the remaining papers, all were screened with a full read, and any further exclusions were noted with reasons. One reviewer (JH) reviewed all references of included papers for additional potential inclusions that may have been missed by the initial search but that otherwise met the inclusion criteria specified above.

### Quality Assessment Methods

A subsequent amendment to the protocol, available on FigShare, describes the quality assessment of the included studies. The studies were assessed using the JBI Critical Appraisal Checklist for Systematic Reviews and Research Synthesis (Institute and Others 2017).

### Extraction, Charting, and Analysis of Data

We extracted data to describe search methods, the types of studies that were included or excluded, the sources of the publications and protocol documents (such as whether the protocol reviewed was sourced by a journal appendix or in a public registry), the endpoints compared (primary, secondary, and similar), any quantitative metrics describing endpoint discrepancies. A quantitative meta-analysis was not planned because of the potential for multiple reviews to contain similar and overlapping datasets without the ability to extract original data. The number and category of endpoint discrepancies, as reported in each paper, were captured, and we calculated ranges, weighted averages (weighted by number of included studies in each paper), and medians. We also conducted a sensitivity analysis, removing data from non-peer-reviewed studies.

The qualitative analysis assessed the methods of each review, including document sources for publications and protocols, and whether protocols originated with a clinical trials registry or with a journal from the researchers themselves. Qualitative data extracted also included reported limitations or weaknesses of the included reviews, implications for policy and future research, and recommendations for reducing outcome reporting bias (ORB) and selective outcome reporting (SOR) in future research.

One author (JH) extracted the following study characteristics into a Google Spreadsheet: Author name(s), publication year, publication ID, publication title, journal of publication, DOI, type of studies reviewed within the review study, number of studies included in the review study, time frame of studies assessed, publication sources of studies assessed, protocol sources of studies assessed, protocol source category (journal, registry, or both), definition of endpoint discrepancy outcome according to the review study, and the number of endpoint discrepancy outcomes found. For each study outcome, the number of studies with endpoint discrepancies found compared to the number of studies assessed in each review was calculated as a percentage.

The database link is accessible via the FigShare project page. Data items that were extracted are also included in a list published in FigShare: https://figshare.com/account/home#/projects/125815.

This study did not require an ethics committee review. The review is reported using the PRISMA-ScR checklist to ensure the completeness of the components expected in a scoping review.

## Results

The initial search in PubMed yielded 546 hits for potentially relevant papers and 84 potentially relevant conference abstracts from ASCO, AACR, and ASH. After title screens, deduplication, and abstract screens, full-text evaluation was performed on eight, with six included according to the criteria (see Figure 1). One of the publications (Permutter) included three parallel research questions and reported results from three comparisons, yielding eight groups of results to report.

**Figure 1:**
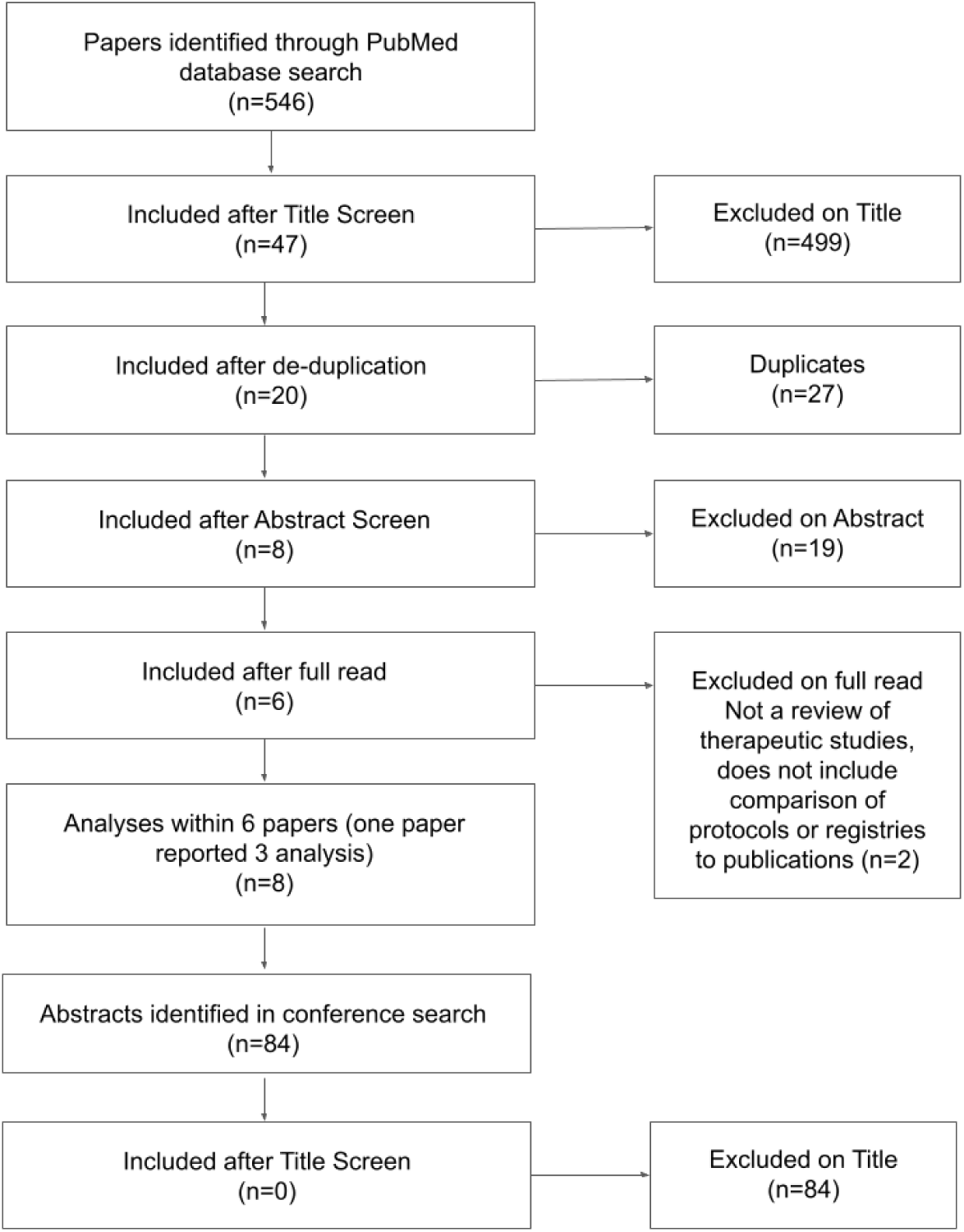
PRISMA Flow Diagram

### Characteristics of Studies

We included six studies published in English-language journals in the US and Europe between 2015 and 2019. Five of the six included data between 2012 and 2015; one included data from 2005 through October 2017 (Aggarwal 2019). Five studies were published as original research papers, and one was published as a letter to the editor (Aggarwal 2019) with extensive supplementary material available online. (See Figure 2)

**Figure 2:**
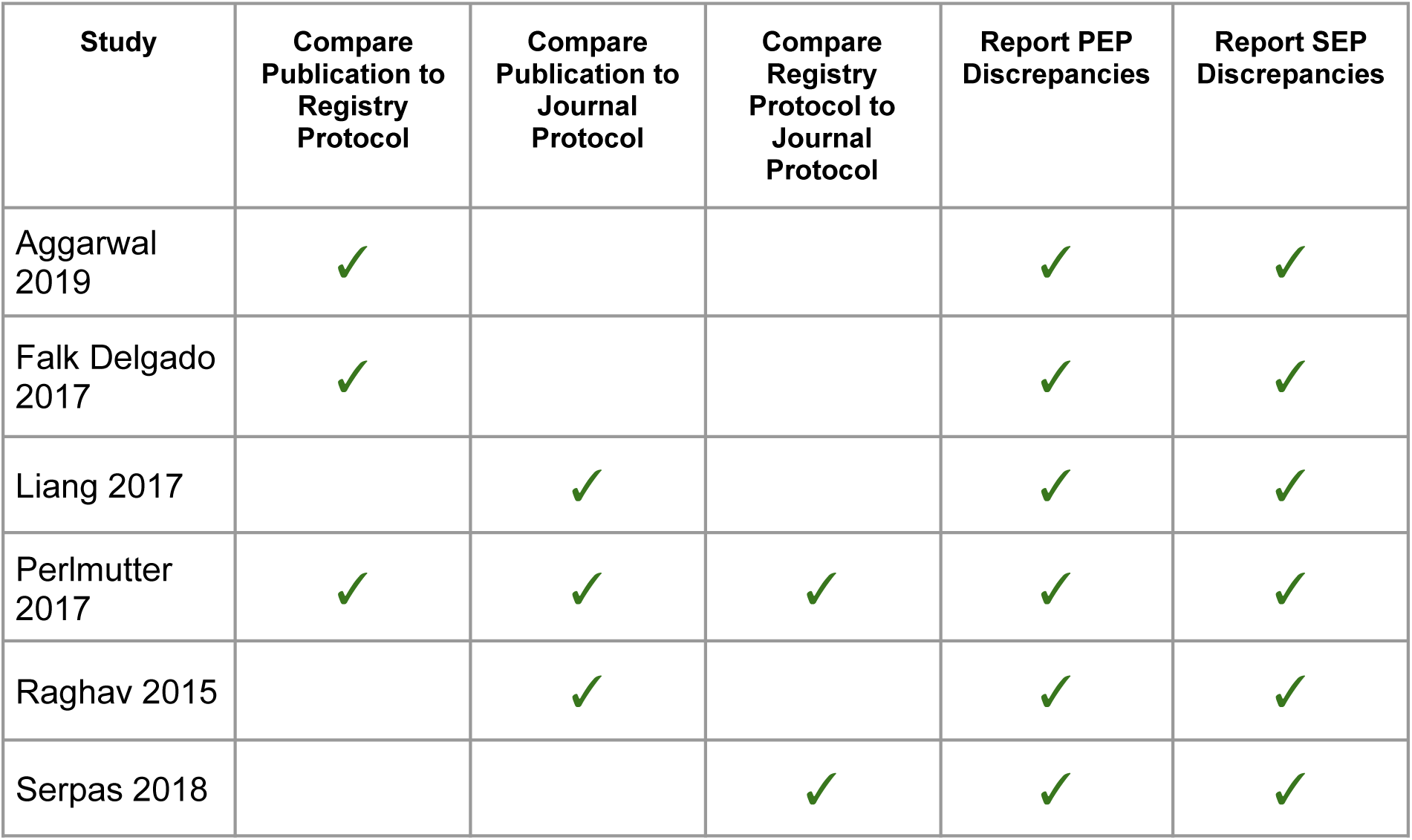
Evidence Map of Study Characteristics

The reviews included between 24 and 217 clinical studies (see Figure 3).

**Figure 3:**
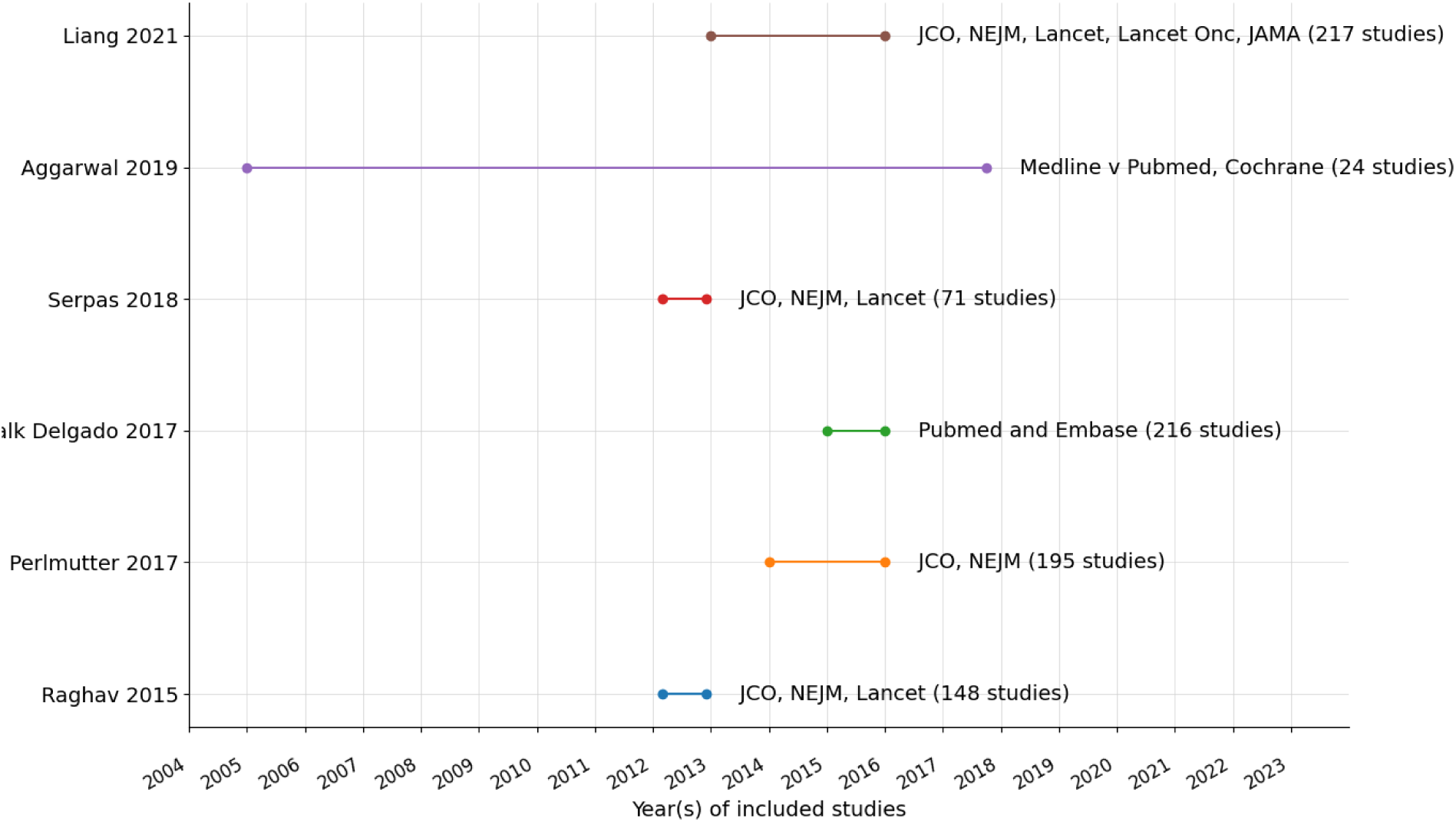
Timeframe and Sources of Papers Assessed by Included Studies

### Quality Assessment & Appraisal

The quality of the included studies was consistently high according to the JBI Critical Appraisal Checklist for Systematic Reviews and Research Synthesis. (Data table in Critical Appraisal tab of this spreadsheet) All studies had “Yes” answers to the applicable questions (7 of 10 items on the JBI instrument). The author notes that more frequently cited quality assessment instruments such as AMSTAR-2 are developed explicitly for the appraisal of systematic reviews of interventional RCTs and, therefore, are not designed to assess systematic reviews of study “metadata” (such as systematic reviews assessing study methods) rather than study outcome data itself.

From a qualitative perspective, the studies that met inclusion criteria had well-articulated research questions, methods, search inclusion and exclusion criteria, and definitions of what constituted Selective Outcome Reporting (SOR) or a similar metric. All reported data sources for the final publications, and all but one reported the data sources for the protocol (for example, whether the protocol was sourced via a clinical trials registry or the journal of publication). Three studies described their limitations, strengths and weaknesses and identified the lack of generalizability beyond oncology. Four of the studies limited their inclusion criteria to the journals that require submission of the protocol.

### Data Sources

Of the five original research papers, three sourced protocols were used to compare to published papers from the journals themselves; two were used for comparison from clinical trial registries, including clinicaltrials.gov and the 14 ICMJE-accepted registries.

All the reviews restricted their inclusion to cancer randomised controlled trials (RCTs); one further restricted to RCTs in Lung Cancer Immunotherapies, and another to RCTs that are primary drug studies reporting on a surrogate primary outcome. Two were restricted to Phase III studies only. Three reviewed trials in specific journals, as the ability to source the full protocol from the journal appendices was a key component of their research methods. For the authors that limited their searches to specific journals, the most restrictive were Permutter et al., who sourced papers only from the *Journal of Clinical Oncology* and the *New England Journal of Medicine*. Raghav et al. and Serpas et al. (two papers using the same dataset) further included *The Lancet*. Liang et al. further included *Lancet Oncology* and the *Journal of the American Medical Association* (JAMA). Aggarwal and Falk Delgado did not restrict their search by journal; both used PubMed to search the Medline database, Aggarwal further used the Cochrane Database, and Falk Delgado further used Embase.

### Definitions of Endpoint Discrepancy / Selective Outcome Reporting

Table 2 shows that events indicating Selective Outcome Reporting or endpoint discrepancy were not defined the same way across the various studies. Variability exists even in the use of the terms “Selective Outcome Reporting” and “Endpoint Discrepancy”, which are at times used interchangeably in the literature. All studies included primary endpoint (PEP) discrepancies, and some also examined reporting of secondary endpoints (SEPs).

**Table 1:** Types of Selective Outcome Reporting discrepancies, their definitions, and potential bias implications.

| Type of SOR | SOR definition | Examples and Potential ORB implications |
| --- | --- | --- |
| Over-reporting | Results or publication report an endpoint that was not pre-specified | New outcome added to support statistical significance or favourable impact/perception |
| Under-reporting | Results or publication fail to report on a pre-specified endpoint | • Subset of analysis not included, and/or<br>• Unfavourable outcome not included |
| Mis-reporting | Endpoint definition or measurement approach shifts between pre-specified endpoint and reported endpoint | • Endpoint measurement or definition changed, and/or<br>• Primary outcome reported as secondary, and/or<br>• Other shift or change that supports a more favourable interpretation of results |

**Table 2:**
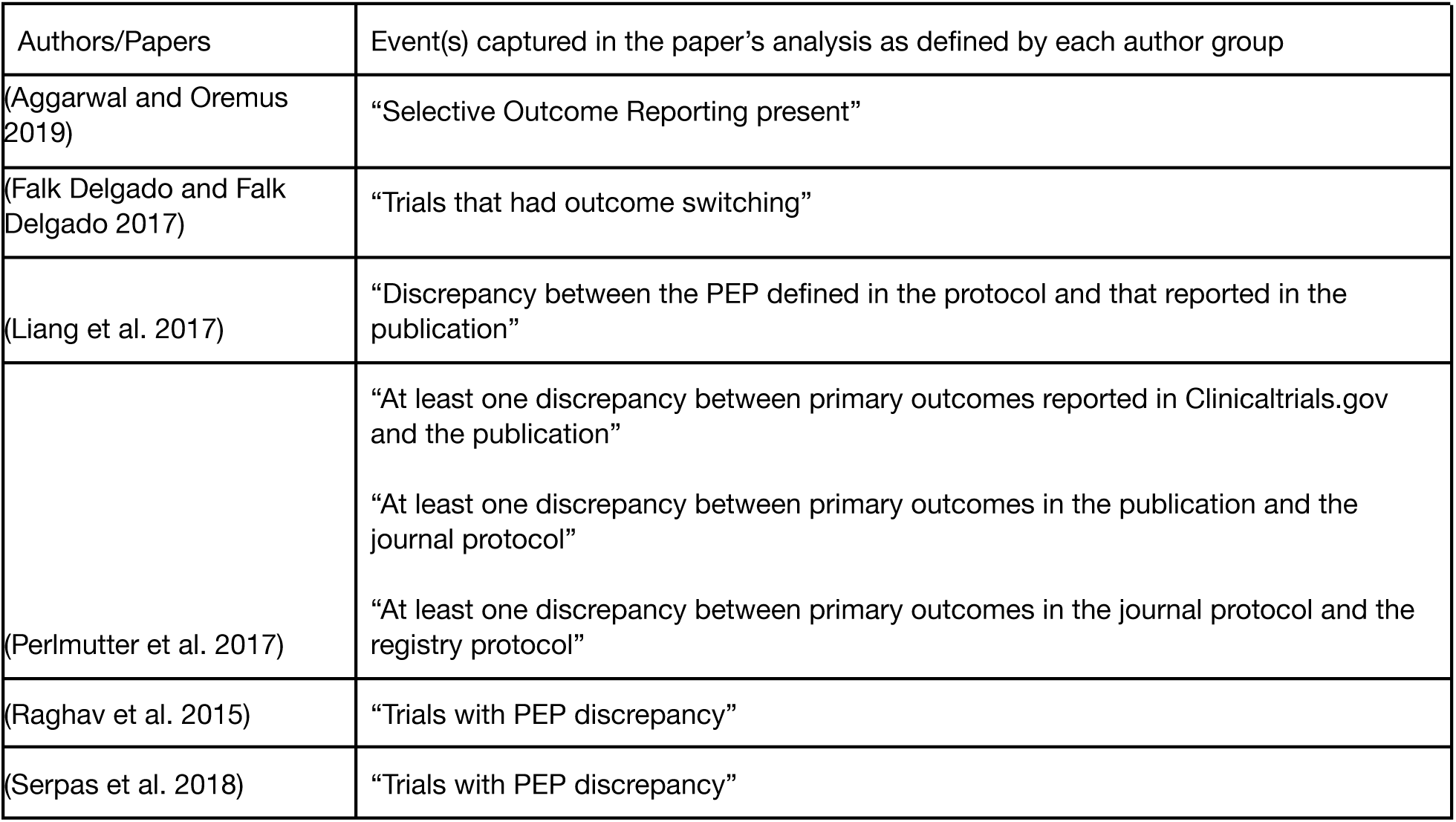
Definitions of “Endpoint discrepancy”.

Table 3 shows that the rate of trials containing endpoint discrepancies between either registry or journal protocol and final publication ranged from 4% to 79%. The median rate of endpoint discrepancy was 15%. Excluding the Aggarwal 2019 letter, which was not peer-reviewed and included only lung cancer immunotherapy RCTs, the range narrowed to 4% to 18%, and the median rate was 12%. A weighted average excluding the Aggarwal letter suggests that about 11% of cancer clinical trials contain endpoint discrepancies.

**Table 3:** Number and characterization of discrepancies found.

| Paper | Events | Studies | % of studies with discrepancies | Definition of event captured | Protocol source | Type of studies included | Primary Endpoint Discrepancy: Category, Number, and Definition | Secondary Endpoint Discrepancy: Category, Number, and Definition | Other or Unspecified Discrepancy: Number and Description |
| --- | --- | --- | --- | --- | --- | --- | --- | --- | --- |
| Aggarwal 2019 | 19 | 24 | 79% | SOR present | Registry | RCTs in Lung Cancer Immunotherapies, 2005-2017 | **Over-Reporting**<br>2<br>Primary Outcome Added<br>**Under-Reporting**<br>4<br>Primary Outcome Omitted<br>**Mis-Reporting**<br>1<br>Primary switched to Secondary | **Over-Reporting**<br>13<br>Outcomes added<br>**Under-Reporting**<br>7<br>Outcomes Omitted<br>**Mis-reporting**<br>3<br>Secondary switched to primary |  |
| Falk Delgado 2017 | 39 | 216 | 18% | Trials that had outcome switching | Registry | Cancer Randomised Controlled Trials (RCTs) that are primary drug studies reporting on a surrogate primary outcome, 2015 |  |  | 20<br>“Major Switching” (PEP addition, switch from SEP to PEP, PEP not congruent with registered PEP or SEP)<br>19 “Minor Switching” (PEP under-reporting, PEP reformulated, PEP selected from several general outcomes in registry) |
| Liang 2017 | 8 | 217 | 4% | Discrepancy between PEP defined in the protocol and PEP reported in the publication | Journals | Phase III cancer RCTs, 2013-2015 | **Under-Reporting**<br>1<br>Omission of Primary Endpoint<br>**Mis-Reporting**<br>6<br>Switching PEP to non-primary | **Mis-Reporting**<br>2<br>Non-PEP switched to PEP | Three publications with endpoint terminology changed from protocol to publication but not deemed to be significant enough to define as a discrepancy. |
| Perlmutter 2017 (Analysis A) | 12 | 65 | 18.5% | At least one discrepancy between primary outcomes reported in Clinicaltrials.gov and the publication | Registry (ClinicalTrials.gov) | Phase III cancer trials in JCO and NEJM, 2014-2015 | **Over-Reporting**<br>3<br>Addition of one or several unplanned PEPs<br>**Mis-reporting**<br>4<br>Change from PEP in registry to SEP in publication |  | **Under-Reporting**<br>5<br>Omission of one or several planned endpoints<br>**Mis-reporting**<br>3<br>Modification of measurement method of one or more outcomes<br>**Other**<br>1<br>Unclear entry in registry preventing assessment |
| Perlmutter 2017 (Analysis B) | 8 | 65 | 12% | At least one discrepancy between PEP in the publication and the protocol | Journals | Phase III cancer trials in JCO and NEJM, 2014-2015 | **Over-Reporting**<br>1<br>Addition of unplanned PEP<br>**Mis-reporting**<br>2<br>Change from PEP in protocol to SEP in publication | **Mis-reporting**<br>1<br>Change from SEP in protocol to PEP in publication | **Under-Reporting**<br>2<br>Omission of planned endpoints<br>**Mis-reporting**<br>6<br>Modification of measurement method of one or several outcomes |
| Perlmutter 2017 (Analysis C) | 12 | 65 | 18.5% | At least one discrepancy between PEP in the protocol and in ClinicalTrials.gov | Journal Protocol vs. Registry Protocol | Phase III cancer trials in JCO and NEJM, 2014-2015 | **Mis-reporting**<br>4<br>Change from PEP registered in clinicaltrials.gov to SEP in the protocol and publication |  |  |
| Raghav 2015 | 9<br>55 | 74<br>74 | 12%<br>74% | Trials with primary endpoint (PEP) discrepancy (comparing journal publication to journal protocol)<br>Trials with non-PEP discrepancies (journal publication to journal protocol) | Journals | Cancer trials published in JCO, NEJM, and The Lancet March-December 2012 | **Over-Reporting**<br>1<br>New PEP<br>**Under-Reporting**<br>1<br>Failure to report planned PEP<br>**Mis-Reporting**<br>7<br>Change in reporting of planned PEP or change in terminology of PEP | **Over-Reporting**<br>28<br>Reports with unplanned endpoints<br>**Under-Reporting**<br>25<br>Studies with >3 planned "non-PEP"s not in report |  |
| Serpas 2018 | 4<br>45 | 71<br>71 | 6%<br>63% | Trials with primary endpoint (PEP) discrepancy (comparing journal **protocol** to **registry**) / **Did not compare publication**<br>Trials with SEP discrepancies | Journal Protocol vs. Registry Protocol | Cancer trials published in JCO, NEJM, and The Lancet March-December 2012 | **Mis-Reporting**<br>4<br>Switch from PEP to SEP, or switch in defined PEP | **Under-Reporting**<br>36<br>Endpoints reported in protocol not included in registry<br>**Over-Reporting**<br>19<br>Registry included SEPs not reported in protocol | **Under-Reporting**<br>25<br>Exploratory endpoints present in protocols not reported in registries |

Figure 4 maps the evidence of endpoint discrepancy comparisons.

**Figure 4:**
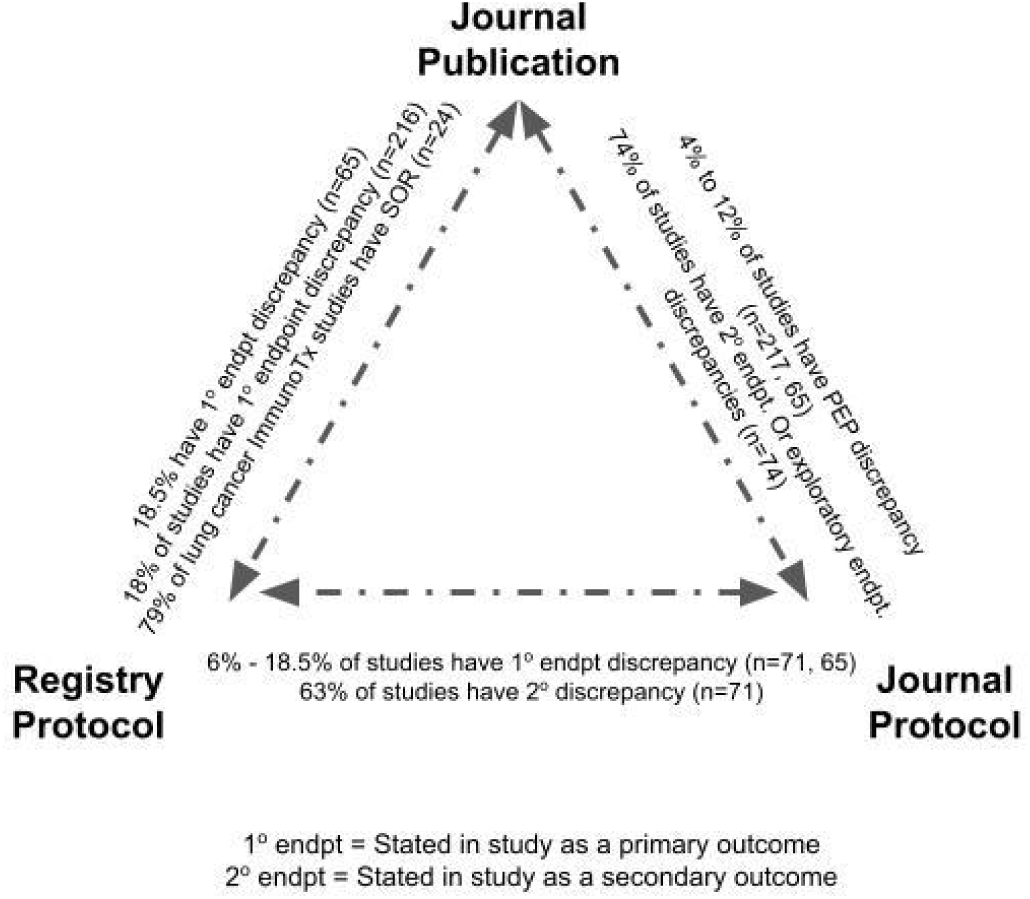
Evidence Map of Endpoint Discrepancy Comparisons

The Raghav paper and the Serpas paper used the same dataset. These papers and the Liang and Perlmutter paper, which includes three analyses, consider both registry-sourced and journal-sourced protocols and report observations regarding the availability and completeness of protocols from both sources. All find that the journal requirement for included protocols is not strictly enforced, as some study papers do not have a corresponding protocol sourced from the journal despite this requirement for publication. As a result, they cannot analyse endpoint discrepancy frequency for trials where the protocol can’t be found. This phenomenon potentially introduces additional bias to the results as researchers who engage in SOR may also be less likely to furnish the protocol to a journal.

Furthermore, Permutter et al. found that endpoint discrepancies occur *between protocol and publication* and also between the protocol *as originally stated* and the protocol *as published in the registry*. The Perlmutter review finds that relying solely on a registry such as clinicaltrials.gov results in “both false-positive identification fo discrepant reporting of outcomes… and false-negative identification of discrepant reporting of outcomes” and further that “only 62% and 59% of trial outcomes described in clinicaltrials.gov contained a description of how and when the outcome would be measured.” Amendment of protocols without subsequent sharing of the updated protocol with all sources, publications, and registries may account for some of this discrepancy. Another factor may be online user interfaces for data entry among registry programs, where word count or character limits may limit the complete entry of all protocol details as originally authored.

## Discussion

### Summary of Main Findings

This paper sought to assess the prevalence of Selective Outcome Reporting (SOR) in cancer studies, understand methods and definitions used to assess SOR, and map evidence, limitations, and gaps. Selective Outcome Reporting (SOR), or endpoint discrepancies, appear to occur in 4% to 79% of cancer clinical study publications, despite standards around pre-specified outcome definitions being articulated more than 25 years ago. From this review, which primarily includes clinical trial papers published between 2012 and 2015 in the most high-profile oncology journals, SOR remains a persistent issue even for studies that can have significant implications for patient care, reimbursement, and resource utilisation. Types of SOR identified by the included studies include addition (over-reporting), omission (under-reporting), and definition change or switching (mis-reporting) of both PEPs and SEPs.

The scope of endpoint discrepancies or SOR in clinical studies described here is consistent with published literature on the topic when top journals are analysed. Kosa et al. 2018 found that 87% of RCTs from 2012 to 2015 in the top 5 journals (200 studies) had an agreement in reporting the primary outcome. Chen et al., 2019 found that one-third of the studies had at least one primary outcome change from among the 389 trials, with outcomes prospectively described in a registry. (Mathieu et al. 2009) indexed 323 RCTs in cardiology, rheumatology, and gastroenterology and found similar results (31%) with “some evidence of discrepancies between the outcomes registered and the outcomes published.”

### Strengths and Weaknesses of this review

This review’s strengths include using a systematic search and appraisal approach and seeking not only to report and quantify rates of SOR in cancer clinical trials but also to characterise gaps in understanding and evidence.

Weaknesses of this review were that the search process and data characterization were completed independently rather than in duplicate. Further, the data extraction process did not use pre-calibrated or tested forms. A further weakness is that the studies included in the review frequently covered overlapping journals and timeframes. Because of this timeframe and journal overlap, some study datasets likely included analysis of similar papers. As the authors of these studies did not list all included studies in their reviews, the full extent and implications of this potential overlap could not be assessed. Additionally, the included studies cover a relatively short time frame, mostly between 2012 and 2015, precluding any assessment of whether the magnitude of Selective Outcome Reporting is changing over time. Two author groups (Aggarwal 2019 and Falk Delgado 2017) hypothesised that due to the extended time frame between when cancer studies commence to when data is published, there would be more likelihood of finding both protocols and papers for more recent studies, as some of the journals selected as search sources only began strictly enforcing the inclusion of protocols in recent years.

Quality assessment of the studies presented a challenge because existing instruments for quality assessment, including the JBI instrument used, have not been developed to assess quality for this type of review. Scoping reviews do not expressly require quality assessment. Because critical appraisal tools for systematic reviews are not designed for quality assessment of studies like the ones included in this scoping review, we may be left with doubts regarding whether all of the identified studies are of high enough quality to warrant inclusion.

For the four included studies that limited their inclusion criteria to the journals requiring protocol submission, this study recognizes that these journals are also among the highest-impact journals in oncology (JCO, NEJM, JAMA, *The Lancet*), resulting in a potential selection bias as these journals are already likely to publish only the most rigorously conducted studies. However, the specificity of the research question and the application to high-impact journals and studies could be a strength in identifying the prevalence of SOR even among studies that are generally well-funded, rigorously monitored, and have significant practice and policy implications due to their frequent use as registrational trials for FDA approvals or indication/label expansions.

Another limitation was that the methods and data reporting of the included reviews did not include the definitions of the over-, under-, or misreported outcomes they found. Therefore, it was not possible to determine if any specific endpoint, such as Progression-Free Survival, Overall Response Rate, Time to Progression, or similar, was more frequently subject to discrepancy than others.

### Implications for Future Research, Suggestions for Improvement, and Unanswered Questions

The lack of a consistent definition of Selective Outcome Reporting suggests a general need for greater consensus on definitions and taxonomy around endpoint discrepancies. The wide quantitative range in the findings reflects gaps in how Selective Outcome Reporting is defined, measured, and reported. The recency of papers that contain SOR further reflects on systemic gaps in the adequate publication of protocols to currently defined standards and regulatory requirements.

This review suggests that SOR is a thorny problem, even within a specific disease area such as oncology. From a policy standpoint, regulators and clinicians should shine a spotlight on this topic to understand how SOR could contribute to biased clinical trial results, whether inadvertent or intentional.

The table below suggests potential ways to improve three of the challenges identified by this review: a lack of consistent language and definition around the problem of SOR itself, discrepancies between protocols and registries that create ambiguity in establishing a baseline for endpoint definitions, and a lack of detail in registry definitions of endpoints that make this most easily accessible, publicly available source less than ideal for assessment of the full SOR picture.

These identified challenges are acknowledged by the authors whose studies were reviewed in this paper. For example, Raghav et al. propose an initiative for Comprehensive Outcomes Reporting in Randomised Evidence (CORRE) to establish a standard for a supplementary section on outcome measures and their statuses to be included with protocol registries; they suggest that this would complement existing protocol drafting guidance such as the Standard Protocol Items: Recommendations for Interventional Trials (SPIRIT) statement and the Standardised Definitions for Efficacy End Points (STEEP) proposal. (Raghav et al. 2015) Furthermore, Raghav et al. and Permutter et al. draw a connection in this way between the problem of SOR and initiatives that drive towards standardisation of endpoint definitions and common outcome measures, such as the COMET initiative. (Williamson and Clarke 2012)

This review found three major research challenges, including suggestions on how to improve or mitigate them.

*Challenge 1: Lack of consistent language around SOR and lack of consistent definitions for what constitutes SOR* No consensus view or standard exists for definitively assessing SOR. This could be mitigated by the use of consistent language and definitions/standards to define over-reporting, under-reporting, and mis-reporting of endpoints/outcomes. For example, the ORBIT study framework on outcome reporting bias (“Welcome to the ORBIT Website” n.d.) provides tools to help systematically assess outcomes reporting bias.

*Challenge 2: Discrepancies are found between “true” study protocols and protocols abstracted to trial registries.* As discrepancies can be found between original study protocols and the abstraction of these protocols in clinical trial registries, possibly due to limited data entry space, amended protocols, or several other reasons, registry protocols may not provide the highest quality information on outcome definitions. A suggestion for improvement would be a requirement for protocols to include organised summaries of all endpoints in a non-redacted, publicly disclosed fashion at the time of study publication to the publishing journal and thus to readers (this approach has been suggested in the CORRE initiative proposal).

*Challenge 3: Lack of detail and specificity in trial registries regarding definitions of endpoint measurement*. Registries frequently contain endpoints that are not defined in sufficient detail to be well understood or accurately replicated. A suggestion for improvement would be requirements for registries to adopt a consistent framework for detail level and quality of outcome descriptions, such as that proposed by Zarin et al. (Zarin et al. 2011) and updated by Perlmutter et al. (Perlmutter et al. 2017) to include domain, specific measurement, specific metric, method of aggregation of data, time frame, the identity of outcome assessor, and blinding of outcome assessor.

### Implications for practice and policy

The frequency with which SOR occurs in oncology studies more than two decades after establishing standards for protocol publication is a concerning signal, particularly as seen in this scoping review where SOR appears in major studies, including registrational trials published in high-profile, high-impact journals. From a policy and practice perspective, an “acceptable” level of SOR has never been established. This study did not illuminate whether the problem of SOR is decreasing over time, nor whether there are guardrails in place at leading journals to reduce the rate of SOR in publication.

For practising clinicians, SOR in key studies could lead to scepticism or negative perceptions regarding the quality of clinical evidence. If clinicians feel they are unable to rely on the rigour of published studies, they may be reluctant to adopt technologies even when these have evidence for patient benefit, as practices such as SOR may erode trust in such evidence.

From a policy perspective, SOR presents challenges to evidence synthesis and developing evidence-based standards such as clinical practice guidelines. Furthermore, as clinical and data science researchers develop tools that use large datasets through patient registries and electronic health records data and then apply machine learning/artificial intelligence algorithms to derive recommendations for practice, population health, or insurance coverage and reimbursement, the presence of SOR points to a potential “garbage in/garbage out” data problem, where definitions of endpoints may be inconsistent but machines interpret them as equivalent. Considering that oncology is one of the focal areas of new clinical development and investment and an area of high spending for public and private insurers globally, policymakers may want to encourage standards and frameworks that reduce/mitigate SOR and strengthen transparency in outcome reporting and definition overall.

## Conclusions

The persistence of Selective Outcome Reporting in cancer clinical trials presents challenges to practitioners and researchers in assessing the quality of evidence. Measuring and reporting on SOR itself is challenging because source documents are sometimes not publicly available or do not fully reflect the study protocol. While mandated standards by regulatory authorities and medical journals have potentially reduced SOR in the past 25 years, they have not achieved full transparency. Additional standards and frameworks, including those that standardise outcome definitions, definition and measurement specifications, and thoroughness of reporting pre-specified and publication outcomes, have been proposed by multiple research groups. Failing to address SOR may lead to erosion of trust in clinical trial results or “garbage in/garbage out” problems in data analytics.

## Data Availability

All data produced in the present study are available upon reasonable request to the corresponding author (Jennifer Hinkel). More information including the study protocol and data are available on the FigShare project page at https://figshare.com/account/home#/projects/125815.

## Acronyms

SOR: Selective Outcomes Reporting
ORB: Outcome Reporting Bias
ICMJE: International Committee of Medical Journal Editors
FDA: US Food & Drug Administration
NIH: US National Institutes of Health
ORBIT: Outcome Reporting Bias in Trials
AMSTAR-2: Assessment of Multiple Systematic Reviews-2
PRISMA-ScR: Preferred Reporting Items for Systematic Reviews and Meta-Analyses - Scoping Review
RCT: Randomised Controlled Trial
JAMA: Journal of the American Medical Association
JBI: Joanna Briggs Institute
NEJM: New England Journal of Medicine
JCO: Journal of Clinical Oncology
ASCO: American Society of Clinical Oncology
AACR: American Association of Cancer Research
ASH: American Society of Hematology
SPIRIT: Standard Protocol Items: Recommendations for Interventional Trials
STEEP: Standardised Definitions for Efficacy End Points
PEP: Primary Endpoint
SEP: Secondary Endpoint

## Contributors

JH contributed to the conceptualisation and methodology of the review, completed the data cleaning, management, and analyses, wrote the original draft, and is the guarantor. CH and CB contributed to the design of the review and writing of the drafts of the manuscript. All authors had full access to all the data in the study and had final responsibility for the decision to submit for publication.

